# Temporal Heart Rhythm Clusters and Physiomorphic Age Mapping: A Deep Learning Approach to Cardiovascular Risk Stratification

**DOI:** 10.1101/2024.04.09.24305561

**Authors:** Gouthamaan Manimaran, Sadasivan Puthusserypady, Helena Dominguez, Jakob E. Bardram

## Abstract

**Purpose:** Understanding the intricate relationships between sleep quality and cardiovascular outcomes can potentially offer new avenues in risk stratification for cardiovascular diseases (CVD). This study aimed to evaluate the significance of biological age predicted through the analysis of sleep stages and nocturnal heart rhythms as a marker for cardiovascular risk.

**Methods:** We leveraged an unsupervised learning approach to generate time-series clusters utilizing whole-night sleep data from *N* = 900 patients, focusing on identifying shifts and consistencies in nocturnal heart rhythms that may indicate variations in cardiac health. Following this, a deep learning model was applied to the time-series clusters to estimate the biological age of the individuals, thereby delineating potential relationships between predicted age, biological age, sleep patterns, and heart rhythms.

**Results:** In a distinct test set of 736 individuals, the predicted age based on this experiment showcased a higher association with mortality (Hazard Ratio (HR) 2.27, p*<*0.05) and CVD risk (HR 3.56, p*<*0.001). Conversely, the age estimated through only nocturnal heart rhythms demonstrated a HR of 2.29 (p*<*0.05) for all-cause mortality and 3.13 (p*<*0.01) for CVD risk.

**Conclusion:** Our findings underscore the high prognostic potential of sleep and electrocardiography data in predicting cardiovascular risks. The method of utilizing predicted biological age derived from sleep stages and nocturnal heart rhythms stands as a significant metric in risk stratification for CVD. Further research in this area might foster novel strategies for early interventions based on sleep quality and cardiac health markers, potentially saving numerous lives through early detection and intervention.

**Author summary:** This study conducted on a large database of sleep data containing physiological signals such as Electrocardiograms, Sleep Stages, anonymized patient information among others shows that the heart behaviour during sleep is indicative of future cardiovascular (CVD) risk and all-cause mortality. This study employs deep learning to predict biological age which is in turn mapped to CVD risk. Through this study, we can see that while heart rhythms during sleep and different stages of sleep (REM, light sleep, etc) does show an association with CVD risk (this exists in previous literature), the more reliable association is found in heart behaviour during specific sleep stages (which is the novelty of our work). We use deep learning to map ECG into different clusters (n=50) using self-supervised learning, and also to find correlation between these clusters and sleep stages while mapping them to their biological age.

## 1 Introduction

Electrocardiogram (ECG) represent a cornerstone in the diagnostic toolkit for Cardiovascular Diseases (CVD), the leading cause of all-cause mortality globally [1]. Leveraging the versatility of ECGs, researchers have utilized it to ascertain a range of critical details including age, gender [2, 3], and historical cardiac events such as Myocardial Infarction (MI) [4], alongside diagnosing cardiac arrhythmias such as Atrial Fibrillation (AFib) and Supraventricular Tachycardia (SVT) [5, 6]. Through the advancements in deep learning, research has now expanded the potential of ECG beyond traditional applications, enabling the deduction of parameters such as sleep apnea [7], ejection fraction [8], body fat percentage [9], etc. These types of approaches may pave the way for deeper insights into a patient’s cardiac health.

Among the mentioned variables, age estimation via deep learning has been a significant area of interest [10]. Despite the crucial role that chronological age plays, offering a straightforward metric denoting the time elapsed since birth, it can be somewhat limited in encapsulating an individual’s health status. This has prompted the exploration of the concept of “biological age”, a potentially more subjective metric that lacks a definitive ground truth. A lot of recent research has embarked on the journey to infer biological ages using various methodologies, each tapping into different dimensions of an individual’s health status - from physical activity levels to chest radiograph assessments [11, 12].

Biological age can be defined as the measure that differs from chronological age by considering not only the time elapsed but also a variety of biological and physiological developmental factors, such as genetics, lifestyle, nutrition, and comorbidities, and is also known as physiological or functional age. [13].

It is imperative to recognize that the biological age inferred from different methodologies might not be the same for a single individual, and it serves as an indicator of specific health aspects represented in the source data rather than a holistic measure of age. Drawing on this, our study delineates two distinctive levels of sleep behavior as mediums to infer biological age:

- **Sleep Stages** such as Rapid Eye Movement (REM) sleep, light and deep sleep
- **Nocturnal Heart Rhythms** acquired from ECG and interpreted through unsupervised clustering

In the subsequent sections, we delve deeper into the rich data landscape available, delineating our approach to harnessing sleep stages-which offer insights into sleep quality, REM cycle durations, etc, and nocturnal heart rhythm behaviors to infer biological age. This exploration holds a mirror to not just the individual’s chronological age but offers a deeper understanding of their cardiovascular health landscape through the lens of sleep-related behavior.

## 2 Methods

This section details the architecture and workflow we use to predict the sleep-related biological age of an individual.

### 2.1 Heart Cluster Mapping

In this study, our objective was to discern the intricate connections between sleep patterns and heart behavior, leveraging an architecture and workflow tailored to predict biological age and its corresponding risk of cardiovascular diseases. Our analysis pivotally relied on the Sleep Heart Health Study (SHHS) dataset [14], which offers electrocardiogram (ECG) data containing the full span of a night’s sleep from the moment of falling asleep to awakening. This dataset presented a vivid tableau of various sleep stages, including REM cycles, light, and deep sleep phases, and instances where patients woke up momentarily, furnishing a robust ground truth for our exploration. We elaborate on this dataset in section 2.3.

#### Data Acquisition and Preprocessing

At the foundation of our analytical pipeline is the preprocessing of the raw ECG signals captured in the SHHS dataset. Our strategy involved dissecting the complex nature of these signals into discernible segments to pave the way for a more nuanced analysis. To this end, we split the continuous ECG recordings into non-overlapping segments, each encapsulating a duration of 10 seconds. It was pivotal to establish a coherent representation of these signals, forming the foundation for the subsequent analytical procedures.

#### Self-Supervised Modelling

In this work, we train our self-supervised model on a separate database which is the PhysioNet 2020 Challenge [15]. Although the details of the self-supervised work is out of the scope of this paper, we introduce the methodology behind training this algorithm to make this paper self-contained.

The input signal is randomly masked 50% and its inverse mask is also applied to the same signal. These two signals are trained in a non-contrastive manner using the cosine similarity loss. Along with this branch, the first masked signal is also reconstructed to its original signal to learn finer details missing inside the signal without the supervision of the second signal. These two paths are trained simultaneously for 25K epochs. The comparison of the performence of this method to BYOL and ResNet is provided in Table 1 below:

**Table 1.** Quantitative Evaluation of our Self Supervised Model

| Method | AUC |
| --- | --- |
| Supervised (ResNet) | 0.89 |
| BYOL | 0.68 |
| Ours | 0.79 |

We compare our method with BYOL where we train the method with all the same parameters and find an 11% increase from our method. We also show the accuracy of a supervised ResNet classifier for a more clear performance comparison.

#### Feature Vector Construction

We then construct a rich feature vector using the self-supervised model mentioned above, thereby mapping the segmented ECG signals into a 128 dimension feature vector. This process aimed to encapsulate the dynamism and intricacies of the heart’s behavior during various sleep stages in a structured digital representation. Through this lens, we envisioned the heart’s nocturnal narrative unfolding in a series of vectors of 128 dimensions, each narrating a 10-second chapter of the heart’s activity during sleep. This step was essential in reducing the data dimensionality while retaining the critical physiological signatures embedded in the ECG signals, furnishing a repository of features primed for deeper analysis.

#### K-Means Clustering and Hyperparameter Tuning

With a rich feature landscape at our disposal, we engaged in the clustering phase to delineate distinctive patterns in the cardiac narratives crafted in the preceding stage. Implementing a K-Means clustering algorithm [16], we explored a range of configurations for the hyperparameter K, ultimately settling at a value of 50. This choice emanated from a balanced approach, seeking to enhance the granularity of the analysis while forestalling an over-complication that could potentially stem from an exceedingly high K value, thereby striking a harmonious balance between detail and computational feasibility.

#### Temporal Heart Vector Construction

Once we had the cluster mapping, we synthesized temporal heart vectors reflecting the heart’s activity trajectory throughout the night for both the training and test cohorts. The visualization of this temporal evolution offered a rich narrative, traversing through a spectrum of 50 potential states, each symbolizing a unique cardiac state, potentially fluctuating through the night as the individual transited between different sleep stages.

At this critical juncture, we crafted visual representations mapping the transitions between different heart states over the course of the night, with a parallel track delineating the various sleep stages. The integration of these narratives into a coherent visualization presented a multifaceted view of the nocturnal cardiac landscape, bridging the physiological narratives with the underlying sleep stages, and laying the groundwork for a deeper exploration into the realms of sleep and cardiovascular health.

### 2.2 Age Regression Track

In our endeavor to ascertain an individual’s biological age as a function of our vital signs cluster vector, we employed three distinct experimental setups, each building on the knowledge and framework established in the preceding section. These setups are detailed as follows:

1. **Utilizing the Heart Cluster Vector alone** (*Heart Cluster model*): The first setup is grounded in the insights gleaned from the heart cluster vector detailed in Section 2.1.
2. **Employing the Sleep Stage Vector** (*Sleep Stage model*): The second experimental configuration exclusively leverages data encapsulated in the sleep stage vector, a detailed record of a person’s sleep stages marked from 0 to 6, illustrating the nuances of each sleep stage and their transitions. This is obtained via the ground-truth annotation of sleep stages in the dataset, but can also be estimated to a reasonable accuracy via EEG and/or ECG data [17, 18].
3. **Correlating Heart Cluster and Sleep Stage Vectors** (*Cluster*×*Sleep model*): Venturing further, our third setup combines the insights drawn from both the heart cluster and sleep stage vectors and their correlation, fostering a rich ground to explore the intricate correlations between cardiac activities and various sleep stages.

Central to these approaches is a cohesive architectural methodology that slightly diversifies when we marry the data streams in the third setup. Here is how we went about it:

#### Mapping Time-Series Data to Learnable Projections

Our raw data comes in time-series categorical format, a structured sequence of data points indexed in time order. To mine this data for biological age predictions, we harnessed it to create learnable projections through embedding layers, a type of neural network layer that learns to map the inputs to a specified dimension space, set here at *N* = 6. This space was envisioned to be a hotbed for learning, holding the potential to unravel complex patterns through learning algorithms.

#### Transforming Vectors for Deep Insights

Post the mapping process, these vectors underwent transformations to suit the neural networks’ analytical requirements, designed to draw deep insights from the sleeping patterns. In scenarios one and two, 2D convolutions were used with convolutions feature sizes of *B* × *T* × *N*. Meanwhile, in the third strategy, we devised a method to interlink the two vectors using 3D convolutions of shape *B* × 2 × *T* × *N*, a rich tapestry holding threads of both heart clusters and sleep stage vectors, ready for deep analysis. In this stage, we also use three parallel blocks of different kernel sizes (5, 15, 31) to find patterns that may be both short and long. We also use the appropriate padding to bring these vectors back to the same sizes.

#### Fine-Tuning and Regression Analysis

The next stage in this process is passing these refined vectors through a shallow 1D convolution network, a structure devised with four distinctive layers, each adding a layer of depth to the analysis, culminating in a pooling stage adaptive to the source vectors’ varying lengths.

Anchoring our approach is the belief in treating age prediction not as a stark classification task but as regression analysis, a mathematical approach attuned to predicting outcomes within a continuous output spectrum. This, we believe, holds the key to understanding the subtle, intricate interplays in the rhythmic data.

### 2.3 Dataset

The Sleep Heart Health Study Dataset [14] is a multi-center longitudinal cohort study to determine cardiovascular and other sleep-disordered breathing. A total of 6,411 men and women aged above 40 years old were enrolled between November 1, 1995, and January 31, 1998. There were two recordings-one at the baseline visit and another 3 years later. Polysomnographic data including but not limited to 1 lead electrocardiogram, Electroencephalogram (EEG), and blood pressure was recorded at these times during the individual’s sleep period. Cardiovascular outcome data were monitored between baseline and 2011.

We use recordings from this dataset that are relevant to our analysis like all night’s ECG data, patient information like age and sex, cardiac outcomes after baseline, and other important information like existing medical conditions like diabetes, hypertension, etc.

In our study, we define a cardiovascular outcome when an individual experiences one or more of the following: Stroke, Chronic Heart Failure, MI, and AFIB. We also only consider patients who do not have any of the mentioned cardiovascular diseases at baseline and assign them as high risk if they experience any of the mentioned outcomes in the following period.

#### 2.3.1 Ethics Statement

The SHHS is a large, multi-center, community-based, prospective cohort study that sought to determine the cardiovascular and other consequences of sleep-disordered breathing (ClinicalTrials.gov Identifier: NCT00005275). The study was performed in accordance with the Helsinki Declaration and each participant provided written informed consent. The current project was approved in April 2020 by the Ethics Committee of National Center of Neurology and Psychiatry (project number: A2020-012). All analyzed data are publicly available (sleepdata.org).

## 3 Results

In this study, we introduce and analyze different estimations of age derived from a variety of biological indicators observed through our series of experiments. The distinct outputs from these experiments provide insights into specific aspects of an individual’s health.

Firstly, the age estimation obtained from sleep stage data offers a glimpse into the quality of an individual’s sleep behavior, effectively quantifying the efficacy and nature of one’s sleep cycle. On the other hand, age estimations drawn from the heart cluster vector experiment depict nocturnal heart activity, offering a distinct yet complimentary lens through which to analyze an individual’s well-being. Though these estimated ages are rooted in divergent biological mechanisms, they do not operate in isolation. Our findings, delineated in Figure 3, suggest that these predictions exhibit a degree of correlation, highlighting a nuanced interrelationship between different facets of biological age. We also find that the chronological age of the patient is correlated with the ages predicted by the *Cluster*×*Sleep* model with an *R*^2^ = 0.295 (*p* = 7.2*e*^−58^) and the *Heart Cluster* model with an *R*^2^ = 0.305 (*p* = 4.5*e*^−60^). We found no significant correlation for the *Sleep Stages* model.

**Fig 1.**
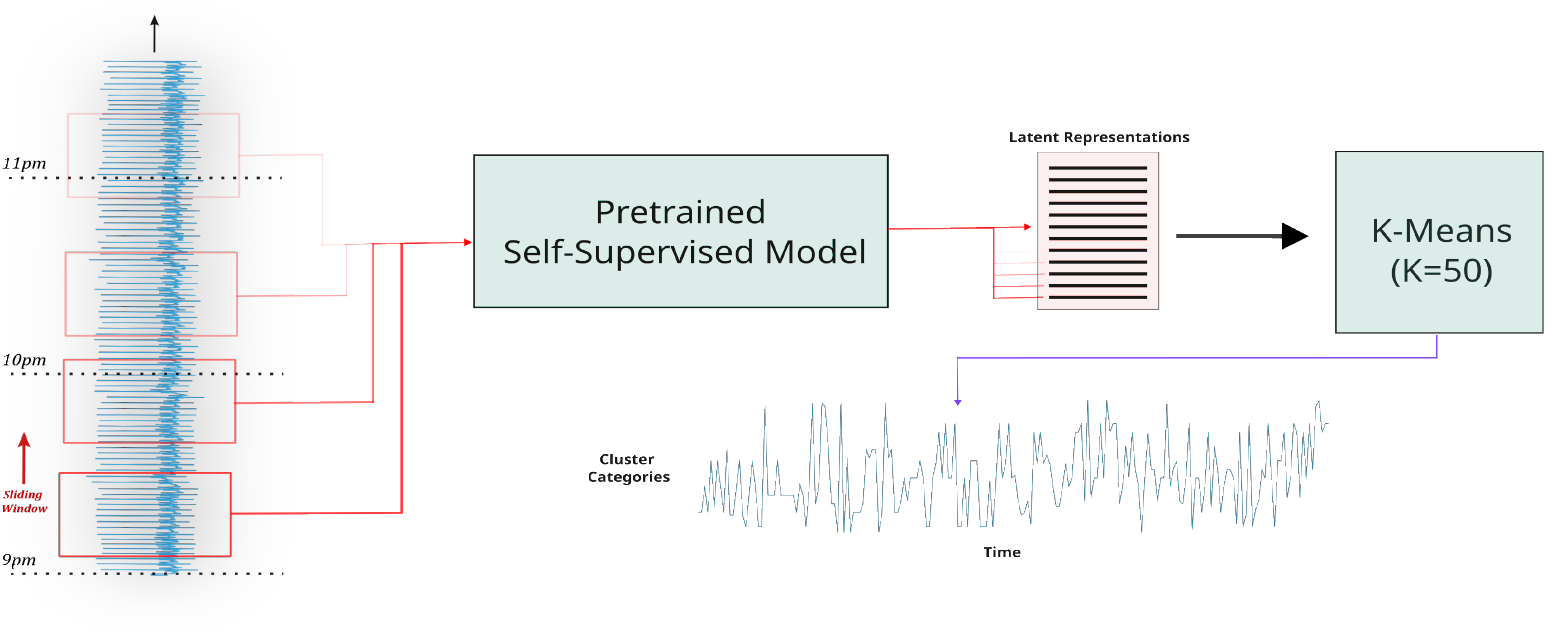
Heart Cluster Vector Formation

**Fig 2.**
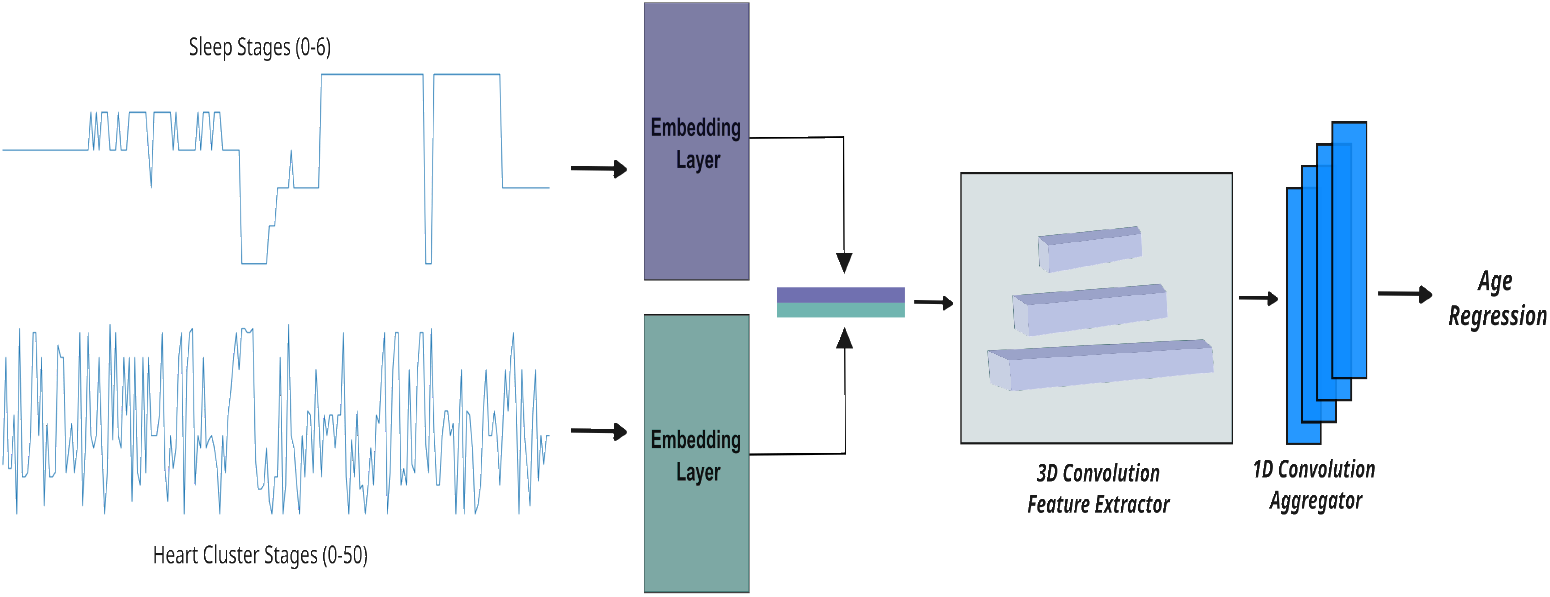
Age Regression Architecture

**Fig 3.**
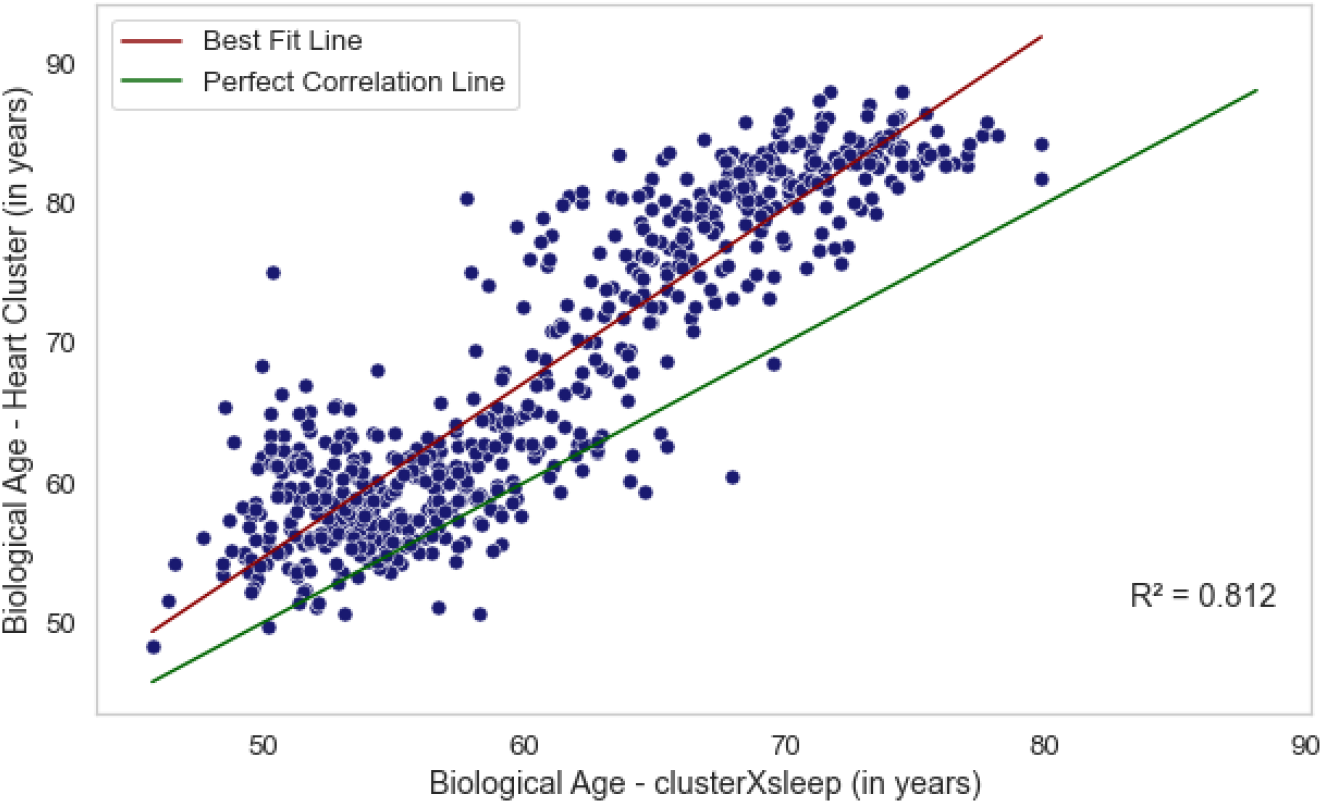
Correlation Plot

To further elucidate the distinctions and correlations between these estimated ages, we segregated the participant pool into differentiated patient groups. Figure 4 showcases box plots that encapsulate the divergences between the chronological age and the predicted ages stemming from all three experimental setups.

**Fig 4.**
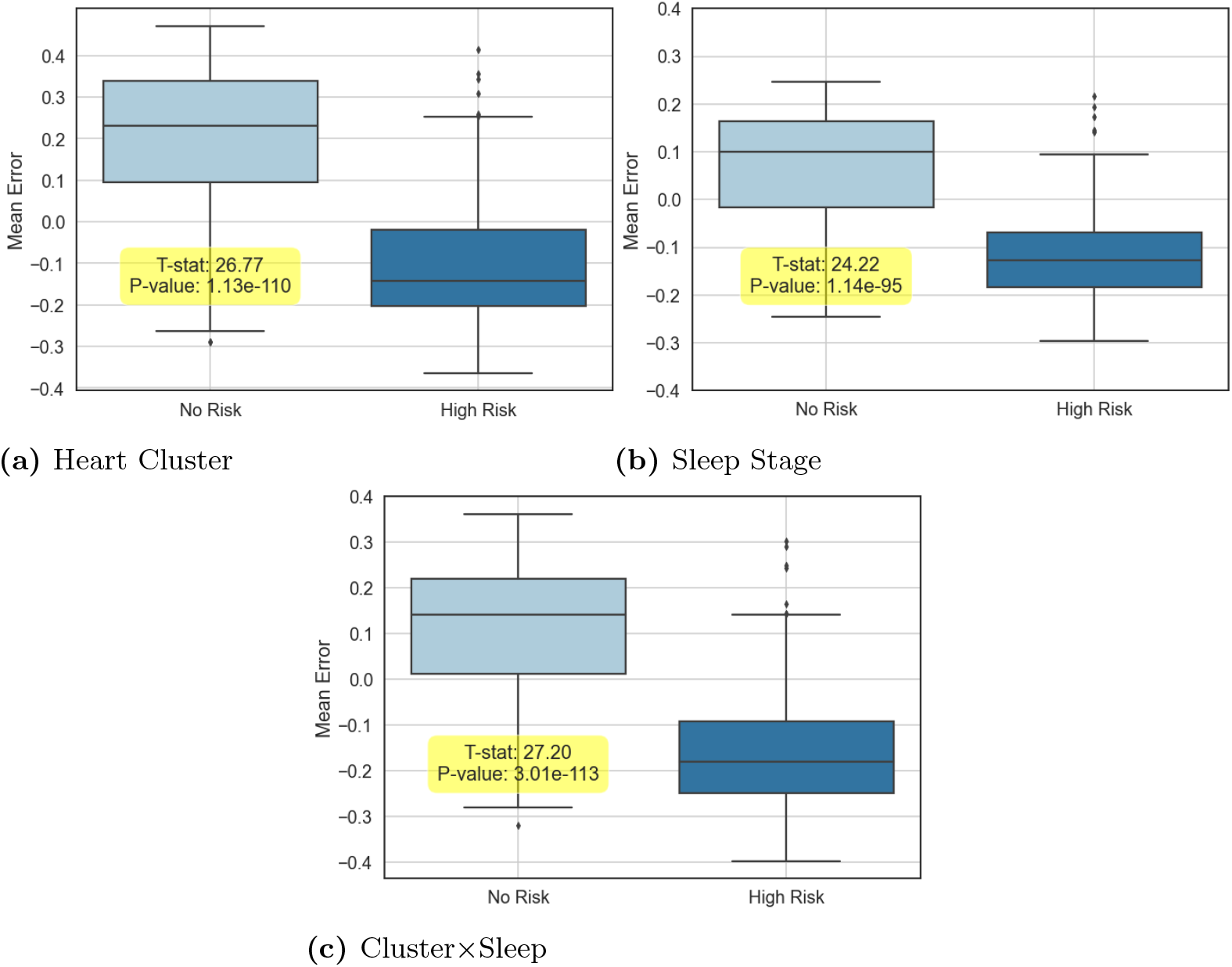
Mean difference across groups for all three experiments for CVD

**Fig 5.**
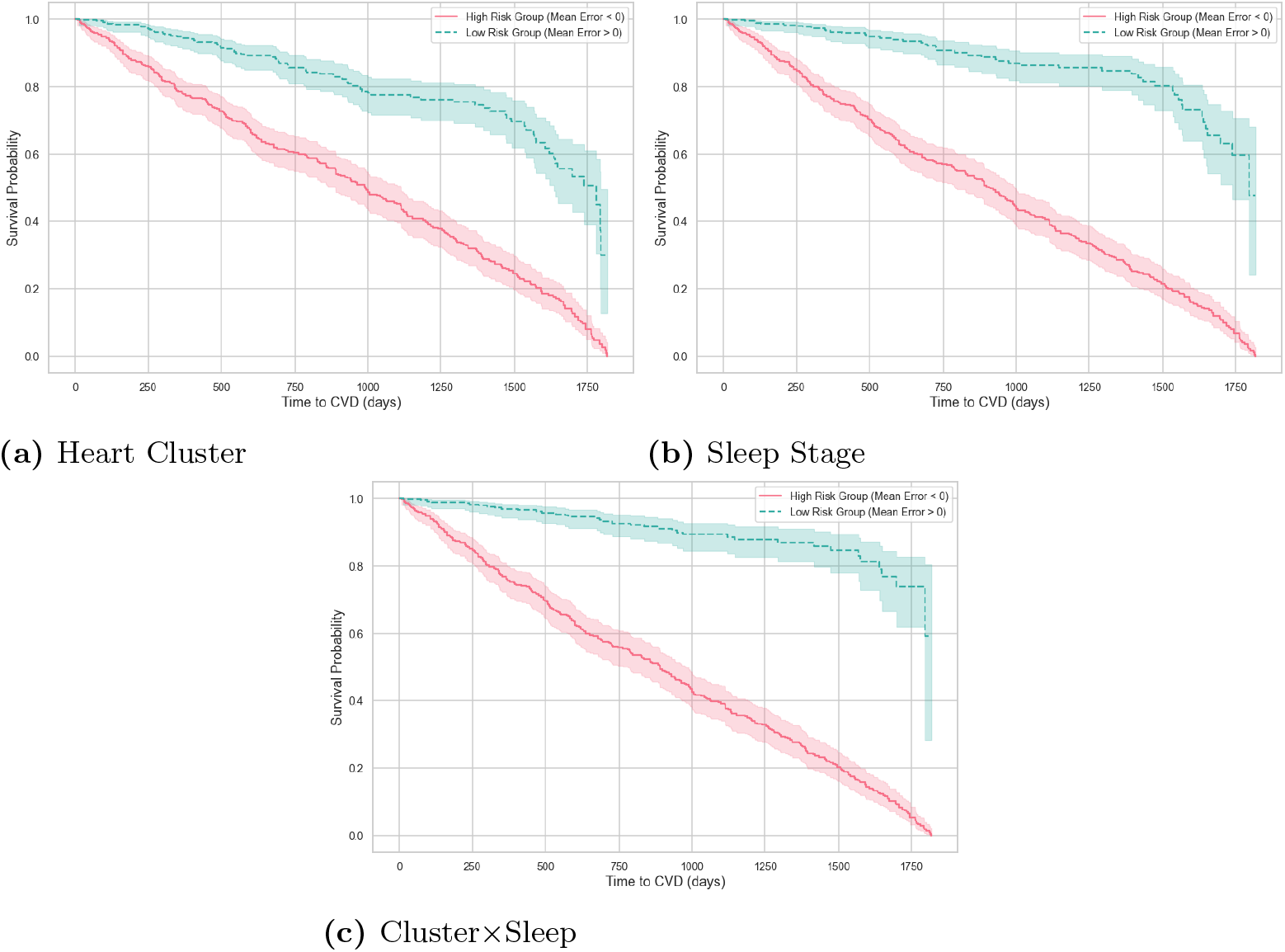
Survival Curves for the three experiments

By aligning these findings with the risk parameters for cardiovascular diseases, a discernible pattern emerges. It is evident that individuals with a heightened risk of CVD consistently exhibit a chronological age that surpasses their predicted age, indicating that our age estimation metrics are attuned to the underlying health trajectories and risk profiles of the individuals. This potentially paves the way for a more nuanced understanding and assessment of health risks anchored in empirically derived age estimations, opening avenues for more personalized healthcare approaches based on individual biological markers.

In each of the three experiments conducted, we delineated the mean disparity as well as the HRs pertaining to all-cause mortality and CVD risk, the details of which are laid out in Table 2. It should be noted that the comprehensive CVD risk encompasses potential hazards stemming from a spectrum of heart-related complications such as strokes, and heart failures, among other ailments detailed in the SHHS database [14].

**Table 2.**
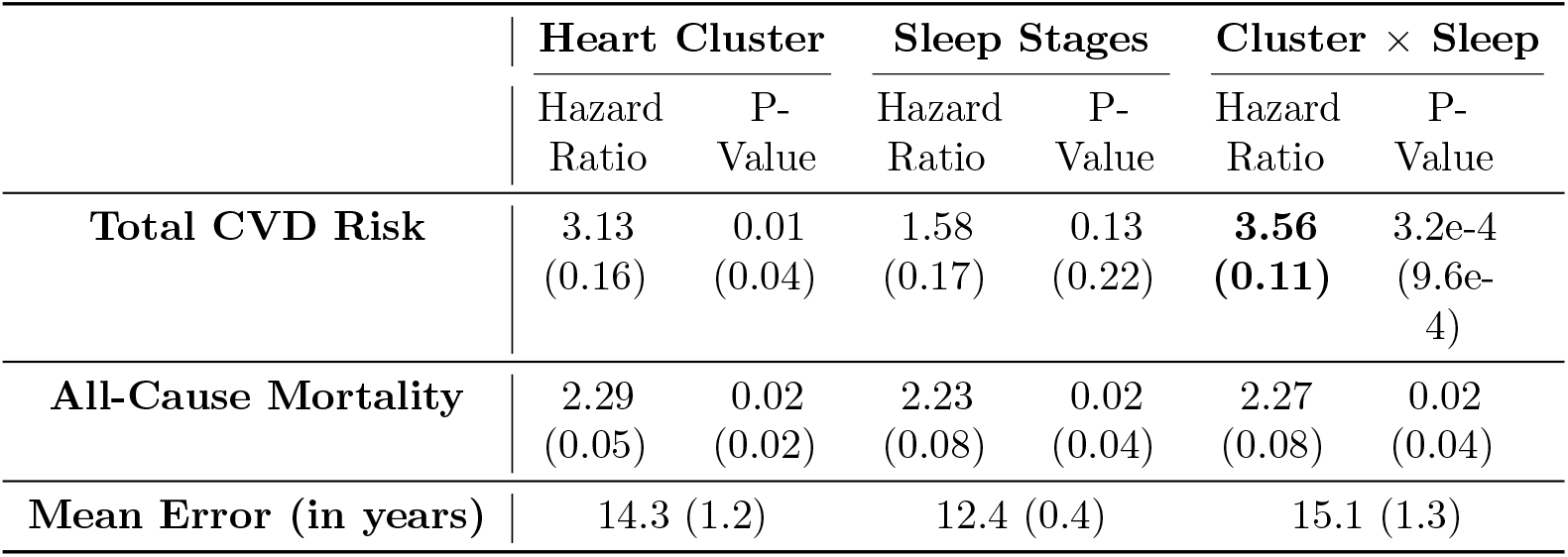
Summary of Risk Stratification of all three experiments. Values are calculated over 10 different seeds and are shown as Mean (Standard Deviation). We show that age computed using the correlation of sleep and heart rhythms gives the most statistically significant result with the highest Hazard Ratio (shown in bold).

|  | Heart Cluster |  | Sleep Stages |  | Cluster × Sleep |  |
| --- | --- | --- | --- | --- | --- | --- |
|  | Hazard Ratio | P-Value | Hazard Ratio | P-Value | Hazard Ratio | P-Value |
| Total CVD Risk | 3.13<br>(0.16) | 0.01<br>(0.04) | 1.58<br>(0.17) | 0.13<br>(0.22) | **3.56**<br>**(0.11)** | 3.2e-4<br>(9.6e-4) |
| All-Cause Mortality | 2.29<br>(0.05) | 0.02<br>(0.02) | 2.23<br>(0.08) | 0.02<br>(0.04) | 2.27<br>(0.08) | 0.02<br>(0.04) |
| Mean Error (in years) | 14.3 (1.2) |  | 12.4 (0.4) |  | 15.1 (1.3) |  |

A critical assessment of the regression setting reveals a substantial mean error as shown in Table 2. However, determining an optimal benchmark for this metric is not straightforward. A scenario where the regression model operates flawlessly, recording a mean error of zero, implies that the predictive power of the model is synonymous with the chronological age in gauging the patient’s risk profile. It is only through the manifestation of a substantial divergence between the predicted and chronological ages that we can foster a metric capable of evaluating risk beyond the limitations of just chronological age.

Therefore, in the grander scheme of the analysis, the mean deviation emanating from the regression model holds a secondary position in importance. The essence of the inquiry is to capture a significant and meaningful difference between the chronological and predicted ages, facilitating a more nuanced understanding of risk, leveraged by parameters beyond mere age. This approach steers us toward a path where risk assessment is a more holistic and intricate endeavor, leveraging a richer set of data and insights.

## 4 Discussion

In this research, we model and assess the deeper correlations between nocturnal heart activity and sleep patterns with the overarching goal of innovating the current methods of stratifying cardiovascular risk. Leveraging the rich data from the SHHS [14] database, we could extract meaningful insights, painting a clearer picture of an individual’s health landscape through our novel age estimation techniques.

The approach of using heart rhythm and sleep stage vectors not only unveiled a rich source of information hidden in nocturnal biological processes but also illustrated the importance of individualized health metrics. The distinction between the predicted age and chronological age brought forth a unique perspective, allowing for a more rounded understanding of an individual’s health and potential risks.

While the correlations found in this study denote a significant step forward in personalized healthcare, it is pertinent to find explainable causes for the differences in the two ages. Though it stands as a limitation in the current study, it opens avenues for further research to refine these models, perhaps by introducing more variables or leveraging more advanced machine learning algorithms to enhance predictive accuracy.

Moreover, understanding that the predicted age carries a meaningful divergence from the chronological age promotes a metric grounded not just in time but in individual biological processes. This represents a shift from a linear perception of age to a multidimensional one, enabling a more granular approach to health risk assessment.

## 5 Conclusion

Through this research, it becomes abundantly clear that the nocturnal sleep and heart rhythm data possesses critical information that holds the potential to transform cardiovascular risk stratification.

We introduced an analytical framework leveraging deep learning models to decipher patterns in time-series data of nocturnal heart rhythms. The emergence of a meaningful difference between the predicted and chronological ages, as unveiled in our research, stands testimony to the potential of this approach in carving out a more individualized, nuanced, and potent tool for risk assessment in cardiovascular healthcare.

The promising results from this endeavor signal a fertile ground for further exploration and refinement of this technique, with a view to constructing a predictive tool that is not just accurate but carries a deep resonance with individual biological rhythms and patterns.

It is our hope that this study acts as a catalyst, encouraging further nuanced research in this direction, and steering the global healthcare community toward a future where risk assessment is not just a function of chronological age but a rich source of individual biological narratives formed through empirical data and innovative analytical techniques.

## Data Availability

The SHHS dataset used in this study has been archived by the National Sleep Research Resource with appropriate de-identification. Permissions and access for these datasets were obtained via the online portal: www.sleepdata.org

https://sleepdata.org/datasets/shhs

## List of Acronyms

CVD: Cardiovascular Diseases
ECG: Electrocardiogram
HR: Hazard Ratio
AFib: Atrial Fibrillation
SVT: Supraventricular Tachycardia
REM: Rapid Eye Movement

## Acknowledgments

This research has been funded by the Innovation Fund Denmark as part of the CATCH project (Project No. #1061-00046B) and the Copenhagen Center for Health Technology. The funder played no role in the analysis or the writing of this manuscript. The Sleep Heart Health Study (SHHS) was supported by National Heart, Lung, and Blood Institute cooperative agreements U01HL53916 (University of California, Davis), U01HL53931 (New York University), U01HL53934 (University of Minnesota), U01HL53937 and U01HL64360 (Johns Hopkins University), U01HL53938 (University of Arizona), U01HL53940 (University of Washington), U01HL53941 (Boston University), and U01HL63463 (Case Western Reserve University). The National Sleep Research Resource was supported by the National Heart, Lung, and Blood Institute (R24 HL114473, 75N92019R002).

